# The Effect of Medical Therapy on Reducing the Risk of Pacing Induced Cardiomyopathy

**DOI:** 10.1101/2024.07.10.24310243

**Authors:** Mahdi S Agha, Christopher L Schaich, Rishi Rikhi, Krupal Hari, George Bodziock, Prashant Bhave

## Abstract

**Background:** High right ventricular (RV) pacing burden can result in pacing-induced cardiomyopathy (PICM).

**Objectives:** To investigate whether ACE inhibitors (ACEI), angiotensin receptor blockers (ARB), and beta blockers (BB) reduce the risk of PICM in patients with high RV pacing burden.

**Methods:** This was a single center, retrospective study which included patients with normal ejection fraction (EF) and complete heart block who underwent pacemaker implantation between 1992 and 2013. The medical therapy group included patients who received ACEI/ARB, BB, or combination of these classes. The control group received neither ACEI/ARB nor BB. The primary endpoint was PICM, defined as upgrade to a biventricular device or reduction in EF to ≤40% without another etiology identified. Fine-Gray subdistribution hazard models accounting for death as a competing risk were used to determine the relationship between medical therapy (ACEI/ARB, BB, or combination) and cumulative incidence of PICM.

**Results:** The study included 642 patients (mean [SD] age 71 [14] years; 51% women). Over 10 years of follow-up, 76 (11.8%) patients received ACEI/ARB therapy only; 49 (7.6%) received BB therapy only; and 86 (13.4%) were exposed to both. PICM occurred in 10 of 211 patients in the medical therapy group (4.7%) and in 30 of 431 in the control group (7.0%). In adjusted analyses weighted for group-switching, the risk of PICM was significantly lower in the medical therapy group compared to the control group (HR 0.59, 95% CI 0.45 – 0.77). Patients exposed to combination therapy had the lowest risk (HR 0.46, 95% CI 0.31 – 0.69).

**Conclusion:** In patients with high RV pacing burden, BB therapy alone or in combination with ACEI/ARBs appears to reduce the risk of PICM within 10 years of pacemaker implantation.

## Introduction

Permanent pacemakers can improve quality of life in patients with symptomatic bradycardia and can be a life-saving treatment in patients with severe atrioventricular conduction system disease. Ventricular pacing is achieved via placement of a pacemaker lead in the right ventricular (RV) apex or septum. Approximately half of patients who receive a pacemaker are dependent on a high percentage of RV pacing.^1^ While the majority of patients are able to tolerate chronic RV pacing, a subset of patients will develop pacing-induced cardiomyopathy (PICM), a form of non-ischemic cardiomyopathy that can progress to clinical heart failure.^1^ PICM can develop at any point after pacemaker implantation.^2,3^ Development of PICM is associated with increased risk of major adverse cardiac events and all-cause mortality.^3^

Currently there is no standardized definition of PICM. It is commonly defined as a reduction in ejection fraction (EF) of ≥10% or a decrease in EF from a normal baseline to ≤40% in the setting of high RV pacing burden and when other etiologies of cardiomyopathy have been excluded.^4^ The reported incidence of PICM varies widely between studies (6 – 39%) depending on the definition used and length of follow-up.^5^

The risk of PICM increases with pacing burden. One study found that each 10% increase in RV pacing burden conferred a 54% increase in relative risk for heart failure hospitalization, and patients who had more than 40% RV pacing burden were 2.6 times more likely to be hospitalized for heart failure.^6^ Meta-analysis has also shown that risk of developing PICM is associated with the presence of conventional risk factors for heart failure, such as male sex, coronary artery disease, chronic kidney disease, and atrial fibrillation.^7^

The treatment for PICM is upgrade to a biventricular pacemaker, known as cardiac resynchronization therapy (CRT). CRT upgrade has been shown to be very effective in improving EF and NYHA class. In one meta-analysis, 85.5% of patients with PICM who received CRT upgrade had an improvement in EF, with a mean improvement of 9.8% in the cohort.^8^ CRT upgrade does not result in a difference in overall mortality.^7^ Conventional guideline-directed medical therapy for heart failure was found to be ineffective at improving EF and NYHA class in patients who have already developed PICM.^2^

While medical therapy has not been shown to reverse PICM^2^, there have been no prior studies investigating whether medical therapy can be effective at preventing PICM. The goal of this study was to investigate whether guideline-directed medical therapy for heart failure – ACE inhibitors (ACEI), angiotensin receptor blockers (ARB), and beta blockers (BB) – are effective at reducing the risk of PICM in patients with normal EF and high RV pacing burden.

## Methods

This was a single-center, retrospective study which included adult patients with a diagnosis of complete heart block who underwent single- or dual-chamber pacemaker with RV lead implantation at Wake Forest Baptist Medical Center between 1992 – 2013. Only patients with a diagnosis of complete heart block were included in the study to ensure all patients had a high RV pacing burden. Patients who received epicardial leads were excluded. Patients with pre-existing cardiomyopathy or reduction in left ventricular systolic function prior to pacemaker implantation were excluded, as well as patients who developed ischemic heart disease during the follow up period. A data extraction software was used to identify patients in the electronic medical record with International Classification of Diseases (ICD-9) diagnosis and Current Procedural Terminology (CPT) procedure codes which fit the study design criteria. The data extraction software returned 645 patients which fit the query parameters. PICM was diagnosed in patients who experienced reduction in EF to ≤40% after pacemaker implantation or underwent upgrade to a CRT device. Manual chart review was performed for patients who developed PICM to verify that there was no alternate cause for the reduction in EF. Three patients were excluded due to an alternate cause for cardiomyopathy.

Patient demographics, medications, and procedures were compiled and analyzed. Patients who had received ACEI, ARB, or BB at any point after pacemaker implantation were included in the medical therapy group. The control group included patients who did not receive ACEI, ARB, or BB at any time after pacemaker implantation. The primary endpoint was the development of PICM. Patients who developed PICM may not have received a CRT upgrade either due to patient preference or high procedural risk. Patients were followed for a maximum of 10 years after pacemaker implantation.

The association of medical therapy exposure with incident PICM was estimated using Fine-Gray subdistribution hazard models with death treated as a competing risk and inverse probability of treatment weighting to account for group crossover during follow-up (i.e., switching from no medical therapy at baseline to any medication exposure within 10 years of pacemaker implantation). Stabilized inverse probability weights conditioned on age, sex, race, and prevalent hypertension, diabetes mellitus, chronic kidney disease, and atrial fibrillation at baseline were generated via logistic regression. Results were additionally adjusted for these covariates in multivariable models. Separate analyses were conducted to examine any medication exposure over follow-up and individual medication groups (ACEI/ARB-only, BB-only, or combination) vs. the control group. We report the subdistribution hazard ratio (SHR) for PICM with 95% confidence intervals (CI). In a sensitivity analysis, we treated group membership as a sequential exposure allowing crossover from the control to medical therapy groups or ACEI/ARB-only or BB-only groups to combination therapy using time-varying Cox proportional hazards models to account for time spent on medical therapy. We further adjusted these models for the covariates described above, and applied stabilized inverse probability weights for death within 10 years of pacemaker implantation conditioned on baseline covariates.

## Results

The study included 642 patients. Baseline characteristics are shown in Table 1. The mean (SD) age of the cohort was 71 (14) years and was 51% female. The medical therapy group included 211 patients (32.9%). ACEI- or ARB-only therapy was prescribed for 76 patients (11.8%), BB only was prescribed for 49 patients (7.6%), and both ACEI/ARB and BB were prescribed for 86 patients (13.4%). In the medical therapy group, 106 patients were on medical therapy at the time of pacemaker implantation, and 105 patients began medical therapy within 10 years after pacemaker implantation. The control group included 431 patients who received no exposure to ACEI, ARB, or BB within 10 years after pacemaker implantation. The median (IQR) follow-up duration was 9 (4 – 10) years. The total incidence of PICM in the cohort was 6.2%. Of patients who developed PICM, 50% underwent upgrade to CRT device. Median (IQR) time to development of PICM was 4.7 (3.2 – 7.1) years. There were 344 deaths (53.6%) that occurred without development of PICM within 10 years of pacemaker implantation.

**Table 1.** Baseline sample characteristics.

|  | **Total Cohort** | **Medical**<br>**Therapy Group** | **Control Group** |
| --- | --- | --- | --- |
| n | 642 | 211 | 431 |
| Age, years, mean (SD) | 71 (14) | 73 (12) | 70 (15) |
| Women | 326 (51) | 115 (55) | 211 (49) |
| Race |  |  |  |
| Black | 68 (11) | 22 (10) | 46 (11) |
| White | 568 (88) | 187 (89) | 381 (88) |
| Other/Not reported | 6 (1) | 2 (1) | 4 (1) |
| Hypertension | 454 (71) | 179 (85) | 275 (64) |
| Diabetes mellitus | 169 (26) | 72 (34) | 97 (23) |
| Chronic kidney disease | 76 (12) | 39 (18) | 37 (9) |
| Atrial fibrillation | 302 (47) | 92 (44) | 210 (48) |
| Medication exposure within 10<br>years of pacemaker implantation |  |  |  |
| ACEI/ARB-only |  | 76 (36) |  |
| Beta blocker-only |  | 49 (23) |  |
| Combination therapy |  | 86 (41) |  |
| Deaths | 344 (54) | 105 (50) | 239 (55) |
| Cumulative incidence of PICM<br>within 10 years of pacemaker<br>implantation | 40 (6) | 10 (5) | 30 (7) |

Data are presented as n (%) unless specified. ACEI, angiotensin converting enzyme inhibitor; ARB, angiotensin receptor blocker; PICM, pacing-induced cardiomyopathy

PICM occurred in 10 out of a total of 211 patients in the medical therapy group (4.7%) and 30 out of a total of 431 in the control group (7.0%). Figures 1 and 2 show the cumulative incidence of PICM over time for the medical therapy group compared to control group and for each medication class compared to the control group, respectively. In unweighted, unadjusted analyses accounting for death as a competing risk, only exposure to combination therapy during follow-up was significantly associated with lower risk of PICM compared to the control group (SHR [95% CI] = 0.64 [0.45, 0.90]; Table 2). For comparison, exposure to ACEI/ARB-only therapy (SHR [95% CI] = 1.02 [0.74, 1.40]) or BB-only therapy (SHR [95% CI] = 0.80 [0.53, 1.22]) were not associated with incidence of PICM (Table 2). Any exposure to either or both medication classes was associated with an unadjusted, unweighted SHR of 0.80 (95% CI: 0.64, 1.01) (Table 2). However, adjusting for baseline covariates revealed that any medication exposure (either or both classes) was associated with significantly lower risk of PICM compared to no exposure (SHR [95% CI] = 0.64 [0.50, 0.82]), an association that appeared to be driven by the BB-only (SHR [95% CI] = 0.61 [0.40, 0.95]) and combination therapy (SHR [95% CI] = 0.53 [0.37, 0.76]) groups (Table 2). Weighting for the probability of switching from no exposure to medical therapy over follow-up did not notably affect results (Table 2).

**Table 2.** Association of exposure to ACEI/ARB, beta blocker, or combination therapy with incidence of PICM over 10 years after pacemaker implantation.

|  | **Subdistribution Hazard Ratio (95% CI) for PICM<sup>a</sup>** |  |  |  |
| --- | --- | --- | --- | --- |
|  | **Unadjusted** | **Weighted<sup>b</sup>**<br>**Unadjusted** | **Adjusted<sup>c</sup>** | **Weighted**<br>**Adjusted<sup>b,c</sup>** |
| No medication exposure | *Ref* | *Ref* | *Ref* | *Ref* |
| Any medication exposure<sup>d</sup> | 0.80 (0.64, 1.01) | 0.80 (0.61, 1.04) | **0.64 (0.50, 0.82)** | **0.59 (0.45, 0.77)** |
| ACEI/ARB-only | 1.02 (0.74, 1.40) | 1.08 (0.78, 1.50) | 0.80 (0.57, 1.11) | 0.79 (0.56, 1.12) |
| Beta blocker-only | 0.80 (0.53, 1.22) | 0.79 (0.52, 1.22) | **0.61 (0.40, 0.95)** | **0.55 (0.34, 0.91)** |
| Combination therapy | **0.64 (0.45, 0.90)** | **0.60 (0.39, 0.92)** | **0.53 (0.37, 0.76)** | **0.46 (0.31, 0.69)** |

ACEI, angiotensin converting enzyme inhibitor; ARB, angiotensin receptor blocker; PICM, pacing-induced cardiomyopathy.
<sup>a</sup>Estimated from Fine-Gray subdistribution hazards models treating death as a competing risk.
<sup>b</sup>Inverse probability-weighted for group crossover during the follow-up period.
<sup>c</sup>Adjusted for baseline age, sex, race, and prevalent hypertension, diabetes, chronic kidney disease, and atrial fibrillation.
<sup>d</sup>Modeled separately from individual groups and includes exposure to any medication class (ACEI, ARB, or beta blocker) within 10 years of pacemaker implantation.

**Figure 1.**
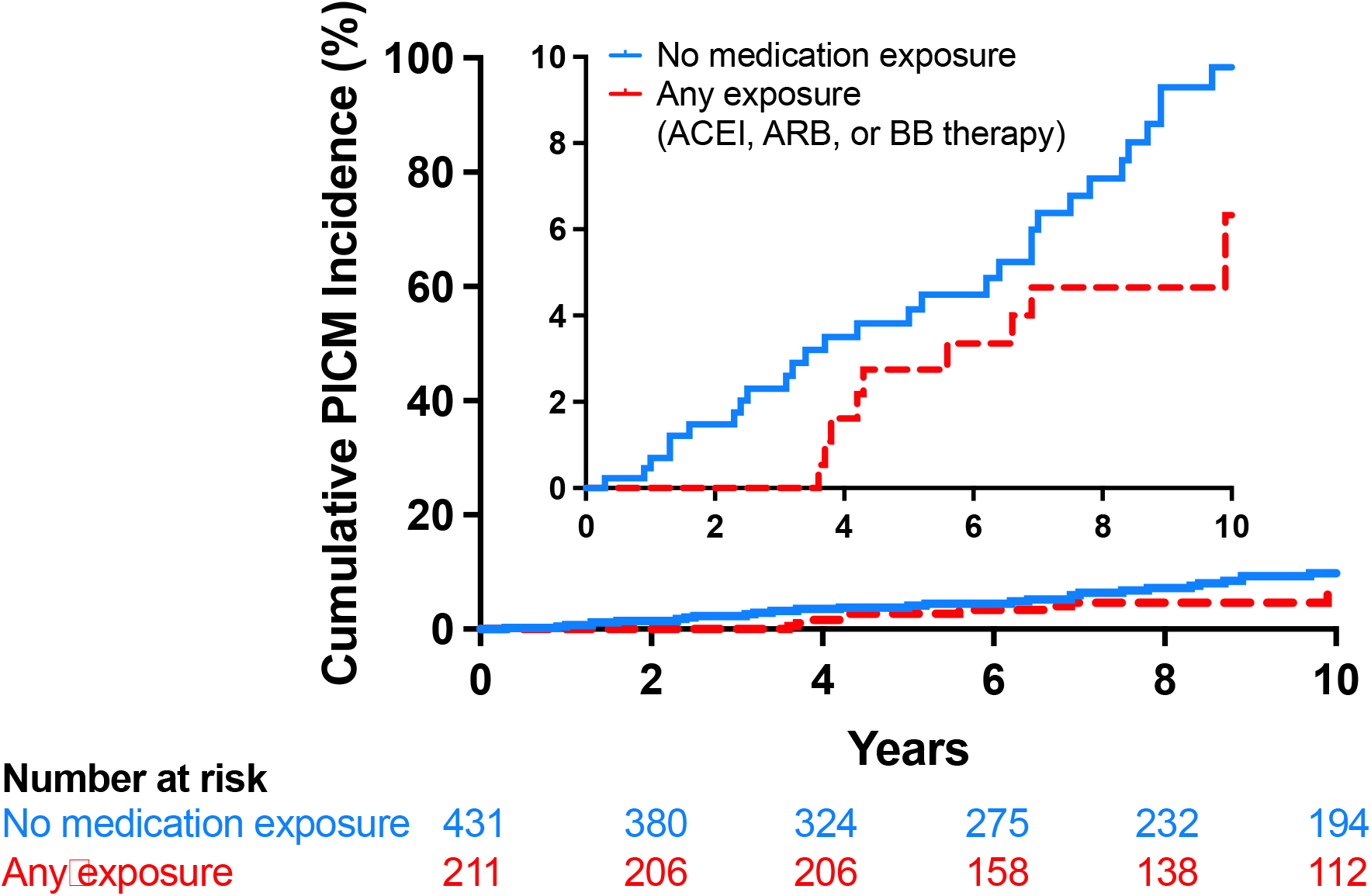
Cumulative 10-year incidence of pacing-induced cardiomyopathy (PICM) after pacemaker implantation among the medical therapy and control groups. ACEI, angiotensin converting enzyme inhibitor; ARB, angiotensin receptor blocker; BB, beta blocker.

**Figure 2.**
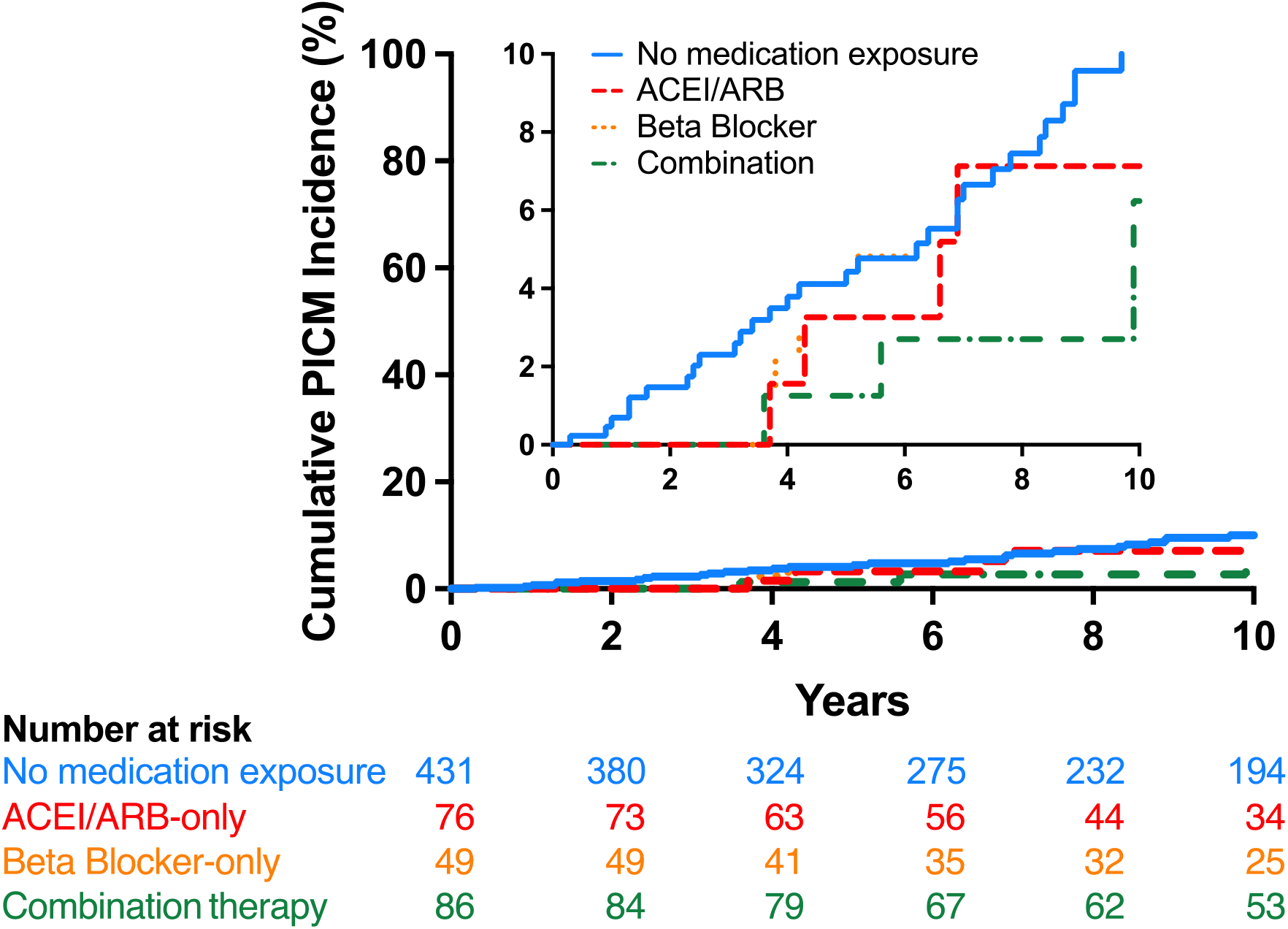
Cumulative 10-year incidence of pacing-induced cardiomyopathy (PICM) after pacemaker implantation among medication subgroups (none, ACEI/ARB-only, beta blocker-only, or combination therapy). ACEI, angiotensin converting enzyme inhibitor; ARB, angiotensin receptor blocker.

Finally, in an analysis treating medication therapy as a sequential, time-varying exposure allowing group crossover, any medication exposure (ACEI/ARB-only, BB-only, or combination therapy) was associated with significantly lower risk of PICM after adjusting for covariates and weighting models for probability of death during the follow-up period (HR [95% CI]: 0.33 [0.13, 0.81]; Table 3). Combination therapy in particular was associated with lower risk of PICM compared to no medication exposure (HR [95% CI] = 0.10 [0.01, 0.78]; Table 3). Point estimates for the ACEI/ARB-only (HR [95% CI] = 0.44 [0.14, 1.33]) and BB-only (HR [95% CI] = 0.48 [0.11, 2.05]) groups in this analysis suggested possible benefit but lacked precision (Table 3).

**Table 3.** Association of sequential exposure to ACEI/ARB, beta blocker, or combination therapy with incidence of PICM over 10 years after pacemaker implantation.

|  | **Time-varying Hazard Ratio (95% CI) for PICM<sup>a</sup>** |  |  |  |
| --- | --- | --- | --- | --- |
|  | **Unadjusted** | **Weighted<sup>b</sup>**<br>**Unadjusted** | **Adjusted<sup>c</sup>** | **Weighted**<br>**Adjusted<sup>b,c</sup>** |
| **No medication exposure** | *Ref* | *Ref* | *Ref* | *Ref* |
| **Any medication exposure<sup>d</sup>** | 0.52 (0.23, 1.18) | 0.45 (0.19, 1.05) | 0.43 (0.18, 1.03) | **0.33 (0.13, 0.81)** |
| **ACEI/ARB-only** | 0.74 (0.26, 2.08) | 0.55 (0.19, 1.63) | 0.61 (0.21, 1.78) | 0.44 (0.14, 1.33) |
| **Beta blocker-only** | 0.52 (0.12, 2.19) | 0.67 (0.16, 2.81) | 0.47 (0.11, 1.97) | 0.48 (0.11, 2.05) |
| **Combination therapy** | 0.24 (0.03, 1.76) | 0.14 (0.02, 1.05) | 0.17 (0.02, 1.44) | **0.10 (0.01, 0.78)** |

ACEI, angiotensin converting enzyme inhibitor; ARB, angiotensin receptor blocker; PICM, pacing-induced cardiomyopathy.
<sup>a</sup>Estimated from Cox proportional hazards models.
<sup>b</sup>Inverse probability-weighted for death within 10 years of pacemaker implantation.
<sup>c</sup>Adjusted for baseline age, sex, race, hypertension status, and prevalent diabetes, chronic kidney disease, and atrial fibrillation.
<sup>d</sup>Modeled separately from individual groups and includes exposure to any medication class (ACEI, ARB, or beta blocker) within 10 years of pacemaker implantation.

## Discussion

In patients with single- or dual-chamber pacemakers who have high RV pacing burden, medical therapy with ACEI, ARB, or BB therapy appeared to reduce the risk of PICM within 10 years of pacemaker implantation. Specifically, patients on BB therapy alone or combination therapy with ACEI or ARB and BB had a lower risk of PICM than patients without exposure to these medications.

The mechanism by which medical therapy may reduce PICM is not clear. BB and ACEI or ARB are known to be beneficial in patients with heart failure with reduced ejection fraction by reducing activation of the sympathetic nervous system and renin-angiotensin-aldosterone system, respectively, which when chronically activated have a negative impact on cardiac function.^9,10^ PICM is a result of the left ventricular dyssynchrony that is produced by RV pacing and usually in the presence of underlying risk factors for cardiomyopathy.^4^ In a healthy conduction system, all regions of the ventricle are depolarized simultaneously to produce a synchronized contraction. Conversely, RV pacing results in slow, dyssynchronous myocyte-to-myocyte depolarization across the ventricle.^11^ Over time, this can result in reduced hemodynamic performance, myofibrillar disarray, adverse cellular remodeling, and decreased global myocardial perfusion.^7,12,13^ Since not every patient with high RV pacing burden will develop PICM, we suspect that patients that develop PICM have underlying susceptibility to non-ischemic cardiomyopathy via chronic, subclinical activation of the renin-angiotensin-aldosterone and sympathetic nervous systems, and thus may benefit from therapy with BB, ACEI, or ARB.

Other methods to prevent PICM in patients with high RV pacing burden have been investigated. One prospective, randomized study showed that implantation of a CRT device in patients with bradycardia and normal EF reduces the risk of PICM when compared to RV pacing.^14^ However, the increased complication rate and cost associated with CRT devices compared to single- or dual-chamber pacemakers prevents this from becoming standard practice. Another strategy to prevent PICM has been conduction system pacing with either His bundle or left bundle area pacing leads. Studies have demonstrated the feasibility and clinical benefits of His bundle pacing, however technical challenges with the procedure and high lead revision rate limit its clinical use.^15^ Left bundle area pacing, a relatively new technique, has shown better lead durability than His area pacing. Long term observational trials have demonstrated that left bundle area pacing preserves EF.^16^ Small, retrospective studies have also show benefit in improving EF in patients who have already developed PICM.^17^ Although prospective randomized trials are still needed, left bundle area pacing is becoming standard practice. Nonetheless, our findings remain relevant to patients with conventional RV leads, patients with failed attempts at left bundle area pacing, and leadless pacemakers, which do not yet or may never have the capability to deliver biventricular or conduction system pacing.

Strengths of this study include its rigorous ascertainment of events, including PICM and deaths, over up to 10-years following pacemaker implantation, and its accounting for death as a competing risk and group crossover during follow-up. Limitations of this study include its retrospective and single-center design rendering causal inferences and generalizability more challenging. Indeed, patients included in this study had close to 100% RV pacing burden due to the presence of complete heart block and thus were at higher risk for developing PICM. Furthermore, our study population had a high prevalence of hypertension, diabetes mellitus, chronic kidney disease, and atrial fibrillation, which are additional risk factors for cardiomyopathy. We speculate that patients who are likely to achieve the most benefit from ACEI, ARB, or BB are those with high RV pacing burdens and traditional risk factors for heart failure. It is unclear whether patients with lower pacing burdens and without traditional risk factors for heart failure would have a similar benefit from medical therapy. Additionally, data extraction software was used to collect information from the medical record and may be prone to inaccuracies, although manual chart review was performed to verify accuracy. Regardless, prospective randomized control trials should be performed to confirm our findings. Finally, assessments of left ventricular function were not standardized due to the study’s retrospective design.

## Conclusions

Patients with pacemakers which do not provide CRT, have a high burden of RV pacing, and traditional risk factors for heart failure are at increased risk for developing PICM. The results of this retrospective, single-center study suggest that medical therapy with ACEI, ARB, or BB, particularly combination therapy with these classes, may reduce the risk of PICM in such patients. Further prospective trials are needed to confirm these results.

## Data Availability

The data that was used for the analysis of this study is available in a de-identified format on request from the corresponding author. The data are not publicly available due to HIPAA and patient privacy.

## Acknowledgments

Special thanks to the support staff at the Clinical and Translational Science Institute at Wake Forest University School of Medicine for their assistance in utilizing using data extraction software to collect clinic data for the project.

## Abbreviations

PICM: pacing induced cardiomyopathy
RV: right ventricle
PPM: permanent pacemaker
CRT: cardiac resynchronization therapy
ACEI: ACE inhibitors
ARB: angiotensin receptor blockers
BB: beta blockers
EF: ejection fraction
AV: atrioventricular

## References

1. Gavaghan C. Pacemaker Induced Cardiomyopathy: An Overview of Current Literature. Curr Cardiol Rev. 2021;18(3):21–26. doi:10.2174/2772432816666210901111616

2. Schwerg M, Dreger H, Poller WC, Dust B, Melzer C. Efficacy of optimal medical therapy and cardiac resynchronization therapy upgrade in patients with pacemaker-induced cardiomyopathy. J Interv Card Electrophysiol. 2015;44(3):289–296. doi:10.1007/s10840-015-0059-4

3. Cho SW, Gwag H Bin, Hwang JK, et al. Clinical features, predictors, and long-term prognosis of pacing-induced cardiomyopathy. Eur J Heart Fail. 2019;21(5):643–651. doi:10.1002/ejhf.1427

4. Merchant FM, Mittal S. Pacing induced cardiomyopathy. J Cardiovasc Electrophysiol. 2020;31(1):286–292. doi:10.1111/jce.14277

5. Kaye G, Ng JY, Ahmed S, Valencia D, Harrop D, Ng ACT. The Prevalence of Pacing-Induced Cardiomyopathy (PICM) in Patients With Long Term Right Ventricular Pacing − Is it a Matter Of Definition? Hear Lung Circ. 2019;28(7):1027–1033. doi:10.1016/j.hlc.2018.05.196

6. Sweeney MO, Hellkamp AS, Ellenbogen KA, et al. Adverse effect of ventricular pacing on heart failure and atrial fibrillation among patients with normal baseline QRS duration in a clinical trial of pacemaker therapy for sinus node dysfunction. Circulation. 2003;107(23):2932–2937. doi:10.1161/01.CIR.0000072769.17295.B1

7. Somma V, Ha FJ, Palmer S, Mohamed U, Agarwal S. Pacing-induced cardiomyopathy: A systematic review and meta-analysis of definition, prevalence, risk factors, and management. Hear Rhythm. 2023;20(2):282–290. doi:10.1016/j.hrthm.2022.09.019

8. Khurshid S, Obeng-Gyimah E, Supple GE, et al. Reversal of Pacing-Induced Cardiomyopathy Following Cardiac Resynchronization Therapy. JACC Clin Electrophysiol. 2018;4(2):168–177. doi:10.1016/j.jacep.2017.10.002

9. Tepper D. Effect of metoprolol CR/XL in chronic heart failure: Metoprolol CR/XL randomised intervention trial in congestive heart failure (MERIT-HF). Congest Hear Fail. 1999;5(4):184–185.

10. Fang JC. Angiotensin-converting enzyme inhibitors or β-blockers in heart failure: Does it matter who goes first? Circulation. 2005;112(16):2380–2382. doi:10.1161/CIRCULATIONAHA.105.586545

11. Cai Q, Ahmad M. Left Ventricular Dyssynchrony by Three-Dimensional Echocardiography: Current Understanding and Potential Future Clinical Applications. Echocardiography. 2015;32(8):1299–1306. doi:10.1111/echo.12965

12. Van Oosterhout MFM, Prinzen FW, Arts T, et al. Asynchronous electrical activation induces asymmetrical hypertrophy of the left ventricular wall. Circulation. 1998;98(6):588–595. doi:10.1161/01.CIR.98.6.588

13. Hohnloser S, Crijns H, van Eickels M, et al. Effect of dronedarone on cardiovascular events in atrial fibrillation. Rev Port Cardiol. 2009;28(3):345–347. doi:10.1097/01.sa.0000360613.90707.27

14. Yu C-M, Chan JY-S, Zhang Q, et al. Biventricular Pacing in Patients with Bradycardia and Normal Ejection Fraction. N Engl J Med. 2009;361(22):2123–2134. doi:10.1056/nejmoa0907555

15. Zhuo W, Zhong X, Liu H, et al. Pacing Characteristics of His Bundle Pacing vs. Left Bundle Branch Pacing: A Systematic Review and Meta-Analysis. Front Cardiovasc Med. 2022;9(March):1–9. doi:10.3389/fcvm.2022.849143

16. Bednarek A, Kiełbasa G, Moskal P, et al. Left bundle branch area pacing prevents pacing induced cardiomyopathy in long-term observation. PACE - Pacing Clin Electrophysiol. 2023;46(7):629–638. doi:10.1111/pace.14707

17. Shan Y, Wang M, Zhang W. the Specific Value of Upgrading To Left Bundle Branch Pacing in Patients With Pacing Induced Cardiomyopathy or Non-Pacing Induced Cardiomyopathy: a Retrospective Study. J Am Coll Cardiol. 2023;81(8):65. doi:10.1016/s0735-1097(23)00509-0

